# Are the Charlson and Elixhauser Comorbidity Indices Reliable Predictors of Postoperative Delirium in Abdominal Surgery?

**DOI:** 10.64898/2026.02.15.26346355

**Authors:** Wesley Chorney, Michael Lisi

## Abstract

**Background:** Postoperative delirium (POD) is a common complication of surgery. It is associated with a number of detrimental effects, including mortality and healthcare costs. We sought to determine whether common comorbidity indices are predictors of POD.

**Methods:** Using the Medical Information Mart for Intensive Care (MIMIC)-IV database, we identified 8022 abdominal surgery procedures across 7212 adult patients. We calculated both the Charlson comorbidity index (CCI) and the Elixhauser comorbidity index (ECI) for each procedure and used logistic regression to predict postoperative delirium, which was defined as delirium within 30 days following the procedure.

**Results:** Models based on either the CCI and ECI were predictive of postoperative delirium (area under the receiver-operator characteristic curve (AUC-ROC) of 0.622 and 0.652, respectively). However, the addition of other factors known to be associated with delirium improved model performance (AUC-ROC of 0.680).

**Conclusions:** Both the CCI and ECI are predictors of postoperative delirium in patients undergoing abdominal surgery. Addition of factors known to be associated with delirium renders additional predictive value and should be included in models that predict postoperative delirium.

## 1. Introduction

Delirium is a common postoperative complication in surgical patients that affects patients, their families and caregivers, and healthcare costs [1]. Delirium is characterized by rapid onset cerebral dysfunction, disorientation, and decreases in cognitive functioning, which follow a fluctuating course. It is related to poorer postoperative outcomes, being associated with postoperative mortality [2], both cognitive and functional decline [3], and increased healthcare burden and costs [4, 5]. In elderly patients, the incidence of postoperative delirium (POD) is high, with estimates ranging from 11.1–45.6% [6]. Delirium is a complex phenomenon, with many interacting predisposing and precipitating factors [7], and it is estimated that a significant portion of cases are preventable [8]. Therefore, the ability to predict patients who may suffer from POD such that they might receive personalized care aimed at reducing their respective risk of POD is an important task.

The Charlson comorbidity index (CCI) was introduced in 1987 [9], with the aim of quantifying comorbidity based on a set of conditions across multiple organ systems. It is known that a CCI of greater than or equal to two is an independent risk factor for the development of POD [10]. A separate meta analysis demonstrated that the CCI was an independent predictor of POD in noncardiac surgery [11]. Additionally, a cohort study demonstrated that it was associated with the development of POD in abdominal and thoracic surgery, specifically in elderly patients [6]. Similarly, the Elixhauser comorbidity index (ECI), introduced in 1998 [12], also quantifies comorbidity in a similar manner to the CCI. Due to its original design, it is used with weightings based on the International Classification of Disease (ICD) coding system used — typically Quan weights for ICD-9 codes [13] or van Walraven weights for ICD-10 codes [14]. The ECI is associated with in-hospital length of stay, hospital charges, and in-hospital mortality [12]. Furthermore, in patients undergoing orthopedic surgery, ECI has been shown to be significantly different in those patients who developed POD versus those who did not [15].

In this study, we investigate to what extent the CCI and ECI serve as predictors of POD in abdominal surgery. In particular, we demonstrate that although these predictors are significantly better than purely random prediction, they have poor discriminatory ability as measured by metrics that are more sensitive to class imbalances. Additionally, we demonstrate that logistic regression can be used in conjunction with these comorbidity indices, as well as other delirium risk factors, to create a better predictive model.

## 2. Materials and Methods

### 2.1 Dataset

We conducted a retrospective cohort study using the Medical Information Mart for Intensive Care IV (MIMIC-IV), version 3.1, a large, single-center, deidentified database containing detailed clinical data for patients admitted to the Beth Israel Deaconess Medical Center between 2008 and 2019 [16]. MIMIC-IV includes demographics, diagnoses, procedures, laboratory measurements, medications, and outcomes for both ward and intensive care unit (ICU) hospitalizations. Use of the database was approved under the data use agreement, and the study was exempt from institutional review board oversight due to the use of fully deidentified data. The database is freely available from PhysioNet [17].

We identified adult patients (18 years of age or more) who underwent general surgical procedures during a hospital admission. Surgical procedures were selected based on ICD-9 and ICD-10 codes, and were chosen to include common general surgery procedures including cholecystectomy, hernia repair, bowel resection, and related abdominal procedures.

For patients with multiple surgical procedures during the same hospitalization, procedures were collapsed to the earliest qualifying surgical date, and the first operation was treated as the index operation. Patients were excluded if there was evidence of pre-existing delirium prior to the index surgery, defined by any delirium diagnosis code recorded before the surgery occurred (as indicated by the timestamp associated with the surgery in the dataset).

The primary exposure was undergoing a qualifying general surgical procedure. Sex and race/ethnicity were obtained from the admissions and patients tables. Baseline comorbidity burden was assessed using the Charlson and Elixhauser comorbidity indices, which were calculated at the time of admission. The ECI was derived from the ICD-10 diagnosis codes using the van Walraven weights [14], and where this was not possible, the ECI was calculated using ICD-9 diagnosis codes using the Quan weights algorithm [13]. Body mass index (BMI) was calculated as weight in kilograms divided by height in meters squared. Height and weight were obtained from the first-day derived anthropometric tables in MIMIC-IV, where available, which provide cleaned and unit-standardized measurements. BMI was treated as a baseline patient characteristic. In addition to these indices and baseline patient information, we also extracted risk factors that are known to be associated with POD, including depression [18], dementia [19], alcohol use [20, 21], and Parkinson’s disease [22].

The primary outcome was the development of postoperative delirium within 30 days of the index surgical procedure. Delirium was identified using diagnosis codes consistent with acute confusional states and delirium syndromes recorded during the postoperative period. Patients with delirium documented prior to surgery were excluded.

### 2.2 Statistical Analysis

All statistical analyses were performed using Python (version 3.13.5). Predictive performance was assessed using logistic regression to transform the CCI and ECI into probabilities, with 10-fold stratified cross-validation. Additionally, a combined logistic regression model is fit using 10-fold cross validation that makes use of the CCI, ECI, patient age, and indicators for whether or not the patient uses alcohol, suffers from depression, dementia, or Parkinson’s disease (which we refer to as the combined model). For each fold, models incorporating either the CCI or ECI as predictors were fit, and performance metrics were computed on the held-out test fold. Fold-specific estimates were then aggregated to obtain overall performance summaries. Model performance was evaluated using area under the receiver operating characteristic curve (AUC-ROC) and area under the precision–recall curve (AUC-PR). The AUC-ROC was used as the primary discrimination metric, given its widespread use and interpretability, while AUC-PR was included as a complementary metric due to the imbalanced nature of postoperative delirium outcomes. For each metric, 95% confidence intervals (CIs) were estimated across the 10 cross-validation folds with the assumption of normality. Mean performance and corresponding confidence intervals are reported. To formally compare the predictive performance of CCI-versus ECI-based models, independent sample two-sided t-tests were used to compare mean AUC-ROC and AUC-PR values across cross-validation folds. For any statistical test, level of significance was set at *α* = 0.05.

Logistic regression models were created using *scikit-learn*, version 1.6.1 [23]. Statistical testing was done with the *scipy* library, version 1.15.3 [24], and visualizations were created with *matplotlib* [25] and *seaborn* [26], versions 3.10.0 and 0.13.2, respectively.

## 3. Results

Table 1 shows the demographics for the cohort used in the study, partitioned by the outcome (postoperative delirium). Where applicable, *p*-values are displayed, using Chi-square tests for binary variables and *t*-tests for continuous variables.

**Table 1.**
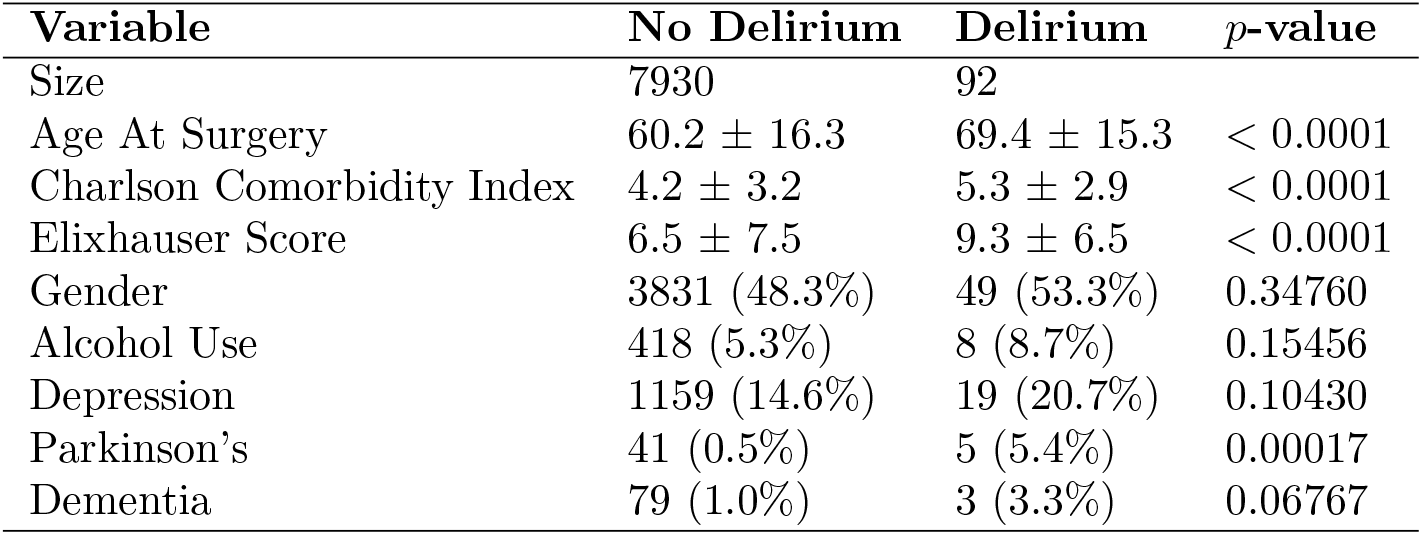
Demographics for the patient cohort used in the study.

The CCI, ECI, and combined models all performed significantly better than a random chance classifier with respect to both AUC-ROC and AUC-PR. The combined model outperformed the CCI-based model (*p* = 0.0412) but not the ECI-based model (*p* = 0.4045) with respect to AUC-ROC. With respect to AUC-PR, the combined model outperformed both the CCI-based and ECI-based models (*p* = 0.0215 and *p* = 0.0107, respectively). Table 2 displays the AUC-ROC and AUC-PR scores for all three models.

**Table 2.** AUC-ROC and AUC-PR scores for the CCI, ECI, and combined models.

| Model | AUC-ROC | AUC-PR |
| --- | --- | --- |
| CCI | $0.662 \pm 0.051$ | $0.023 \pm 0.015$ |
| ECI | $0.652 \pm 0.080$ | $0.020 \pm 0.008$ |
| Combined | **$0.680 \pm 0.066$** | **$0.060 \pm 0.044$** |

Figure 1 displays the observed delirium risk against the quintiles of predicted risk for each model. The combined model shows the desired behaviour — a monotonically increasing function across increasing quintiles. Both the CCI- and ECI-based models are somewhat more volatile, and predicted risk does not uniformly increase as observed delirium per quintile does. For instance, the highest quintile of predicted risk based on the CCI-based model has significantly less POD than the previous quintile. This behaviour is similar in the ECI-based model, but in both the second and final quintiles.

**Figure 1.**
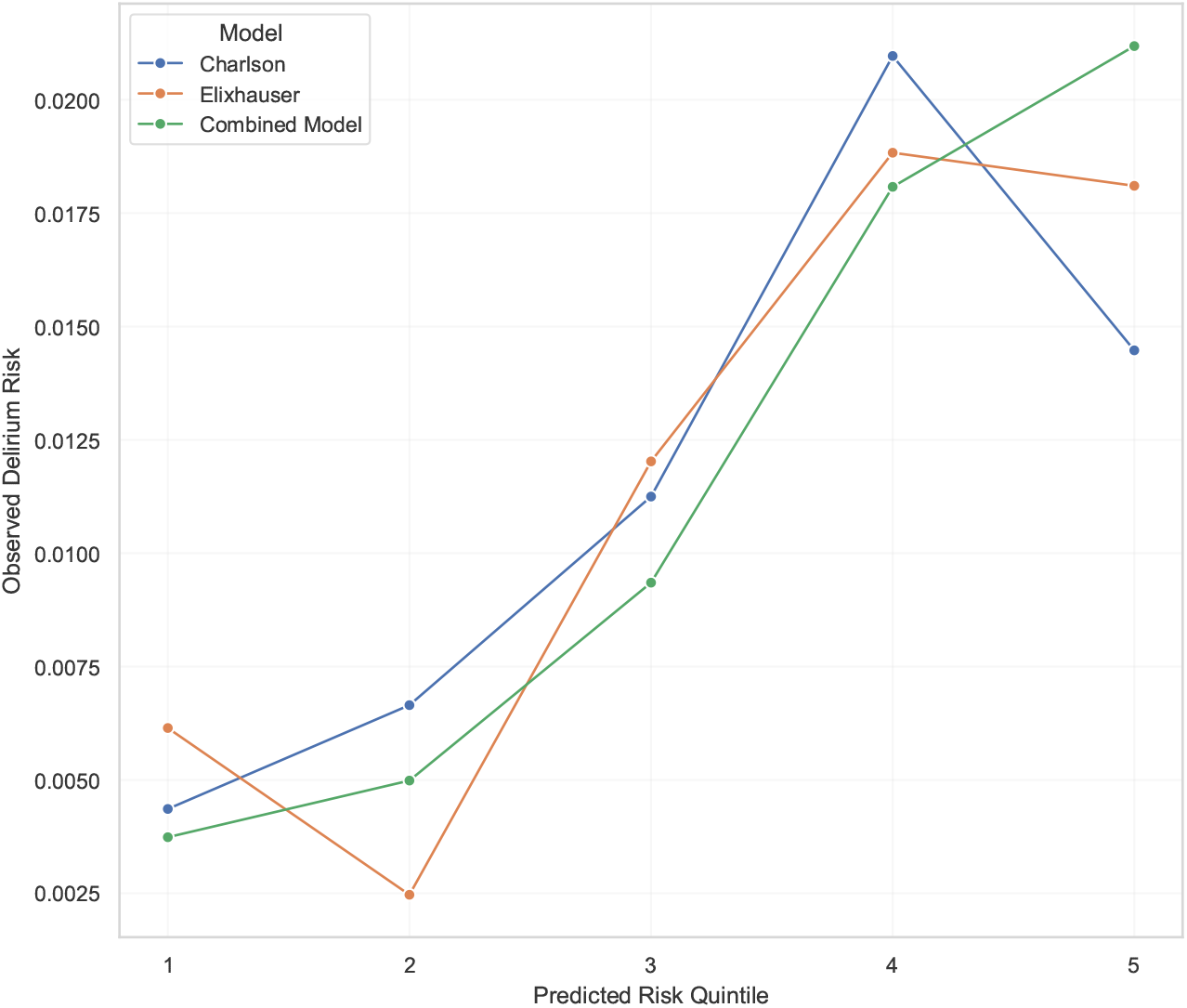
Delirium incidence by risk score quintiles for the CCI, ECI, and combined model.

Figures 2 and 3 show the ROC and PR curves for each model, as well as displaying the area under the curve for each model. While the AUC-ROC is a much more common metric, the AUC-PR discriminates better between model performance in the case of class imbalance [27]. We observe that the combined model has the best mean performance with respect to both AUC-ROC and AUC-PR, performing significantly better than both models with respect to AUC-PR.

**Figure 2.**
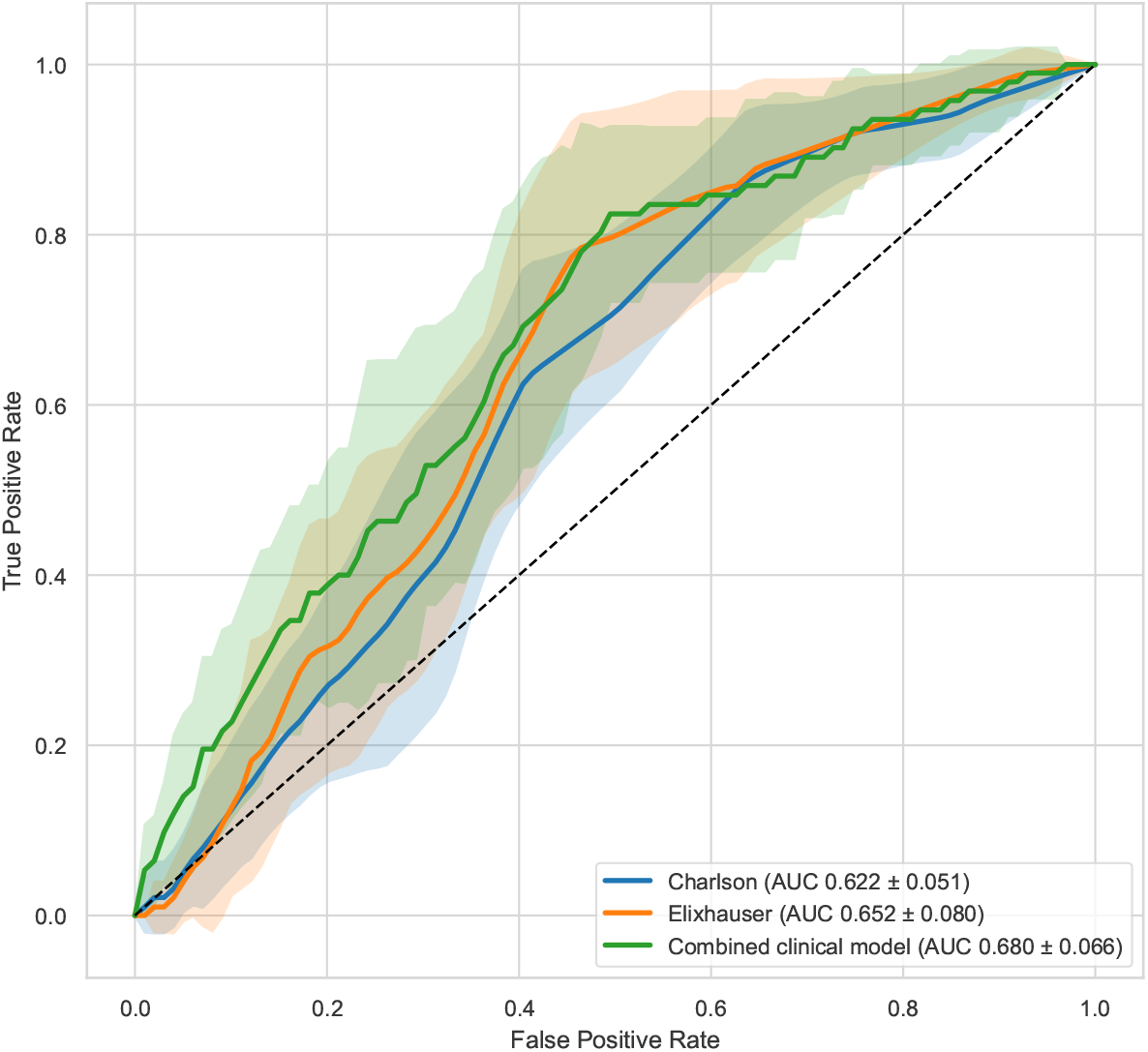
AUC-ROC scores and the ROC curves for the CCI, ECI, and combined model.

**Figure 3.**
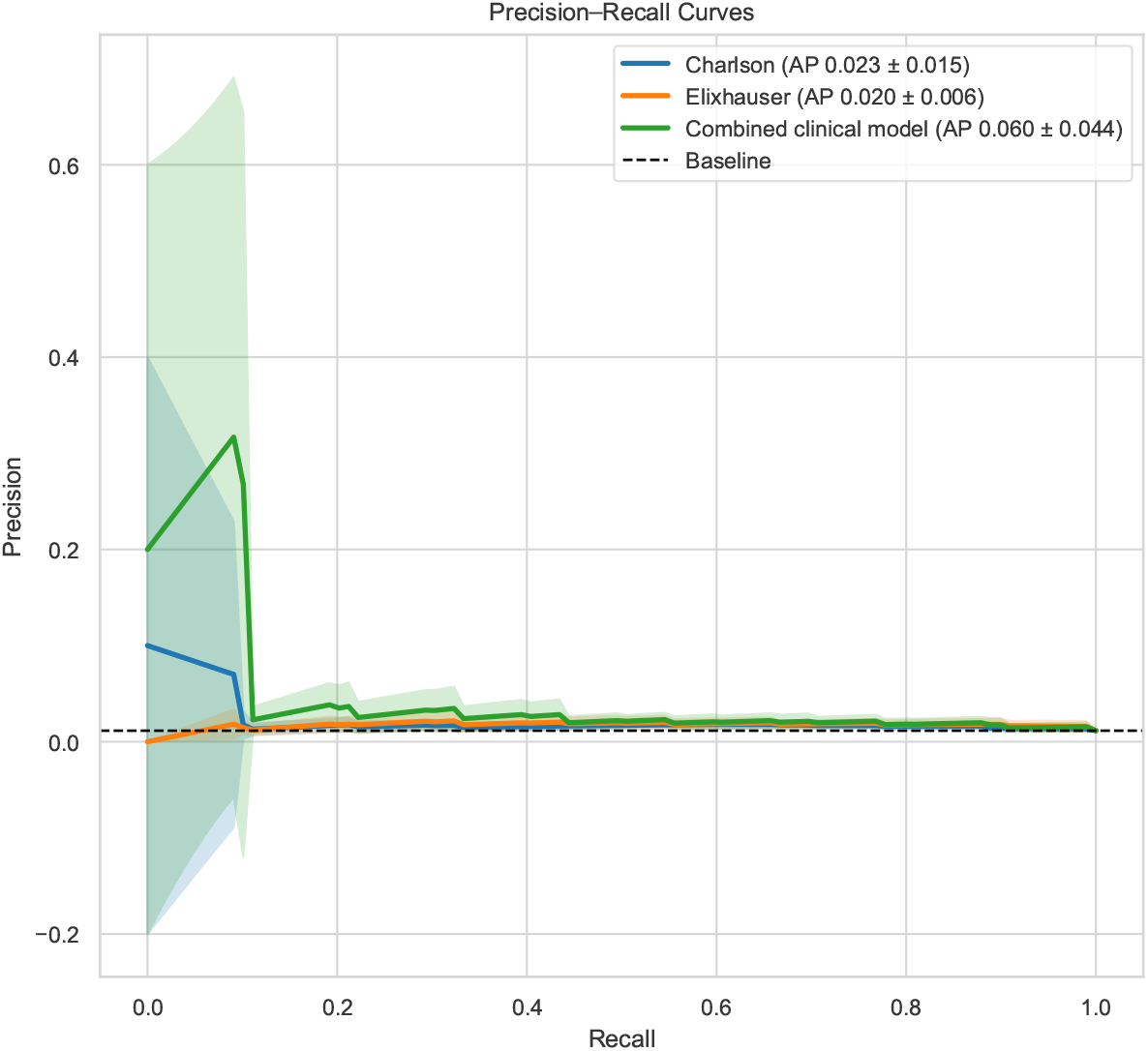
AUC-PR scores and the precision-recall curves for the CCI, ECI, and combined model.

Finally, Figure 4 shows violin plots for the CCI, ECI, and combined models. This figure demonstrates, for each model, the distribution of predicted probability of POD separated by actual outcome. The combined model achieves better discrimination between those patients who experienced POD versus those that did not, while both the CCI- and ECI-based models have very similar predicted POD risk between the two outcomes.

**Figure 4.**
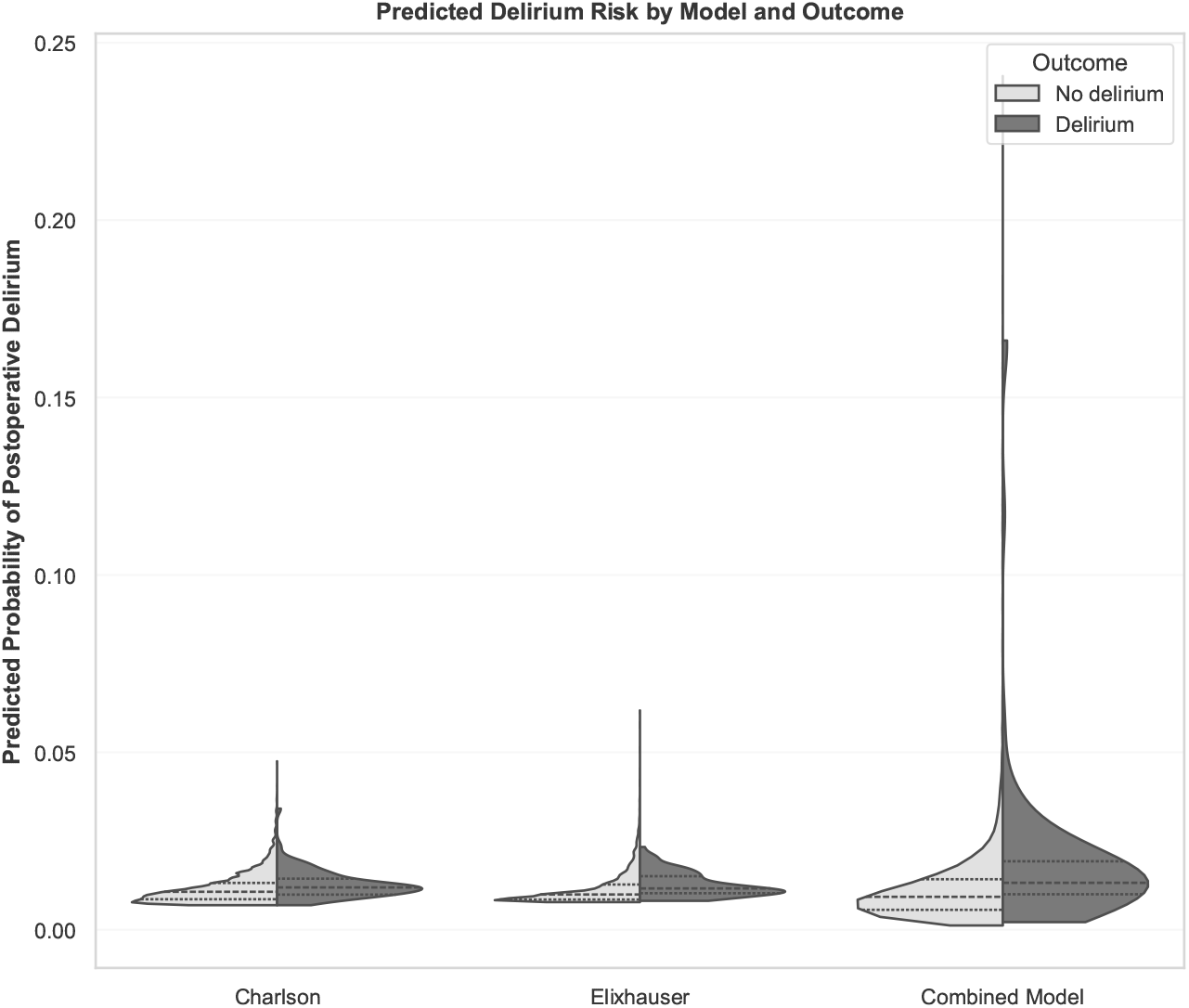
Violin plots for the CCI, ECI, and combined model.

## 4. Discussion

Overall, we demonstrated that both the Charlson and Elixhauser comorbidity indices are predictors of postoperative delirium in abdominal surgery. Notably, this is true even in a cohort consisting of adult (not purely elderly) patients. While this confirms that both indices carry prognostic information for POD, we also demonstrated that a combined model, consisting of both indices as well as other risk factors known to be associated with POD, performs significantly better than models containing one of the indices alone. Therefore, while our study confirms that the CCI and ECI are relevant factors in the prediction of POD in the case of abdominal surgery, it also highlights that additional factors should be evaluated when predicting postoperative delirium. Moreover, any additional factor shown to be associated with POD in such a model can then be optimized prior to surgery, with the goals of reducing the incidence of POD.

While the CCI and ECI are straightforward to calculate (for instance, the ECI requires only administrative-level data), they omit many factors known to be associated with POD. Therefore, while they are of clear clinical value with respect to their association with mortality [28], and despite their clear association with POD, it is important to include additional variables in any model that seeks to predict POD. We note that the combined model significantly outperforms CCI- and ECI-based models with respect to AUC-PR, which is a sensitive metric for imbalanced class prediction problems. This is due to the inclusion of additional factors that are linked to POD. We note that it would be possible to develop a more extensive model with better discrimination between patients who will suffer from POD versus those who do not; however, this was not the main goal of this study. We extended previous work to demonstrate that CCI and ECI are valid predictors of POD in abdominal surgery in an adult (not only elderly) population.

The advantages of the current study lie mainly in its generalizability. The cohort obtained from the MIMIC-IV database consists of 8022 procedures across 7212 patients, with a wide range of ages and covering common procedures in abdominal surgery. The cohort is not restricted to only elderly patients, nor is it restricted to a single comorbidity index. Additionally, we show that both comorbidity indices are valid predictors of POD for metrics that are more sensitive to prediction problems involving rare outcomes. Furthermore, we demonstrated the importance of including additional factors in a model for predicting POD. However, there are some limitations worth mentioning. The MIMIC-IV database consists of data from patients who visited the emergency department or were admitted to the ICU in the Beth Israel Deaconess Medical Center. Therefore, the cohort is most likely biased towards sicker patients (for instance, patients who were hospitalized but never admitted to the ICU or never visited the emergency department would not be included). Furthermore, the cohort was defined by ICD-9 and ICD-10 codes for procedures in abdominal surgery and for the diagnosis of delirium. Therefore, the cohort is also influenced by the accuracy with which these codes were used at the Beth Israel Deaconess Medical Center. Finally, the patients are also drawn from a single center, therefore, the results may not generalize to a patient population that differs significantly from the population from which the cohort was drawn.

While the CCI and ECI are the most commonly used comorbidity indices, other comorbidity indices exist, such as the Johns Hopkins Aggregated Diagnosis Groups [29] or the Wright-Khan index [30]. Further work could investigate whether these less used comorbidity indices carry more prognostic value with respect to POD in abdominal surgery patients. Additionally, while we showed that addition of factors associated with POD significantly strengthened the logistic regression model, this was by no means an exhaustive effort. Future studies could include hundreds of potential factors and use stepwise logistic regression to determine which collection of factors are independent predictors of POD to create a more performant model. Furthermore, different models could be used. While we focused on logistic regression, due to its convenient statistical interpretability, more advanced machine learning models could be investigated and could optionally be combined with methods such as Shapley values [31] to confer explainability.

## 5. Conclusion

We conducted a retrospective cohort study to determine to what extent the CCI and ECI were predictors of POD in a broad cohort of patients undergoing abdominal surgery. Both the CCI and ECI were confirmed to be valid predictors of POD; however, inclusion of factors known to be associated with POD (age, alcohol use, dementia, depression, and Parkinson’s disease) significantly improved prediction of POD. We note that while both the CCI and ECI are predictive of POD, it is important to include other factors to achieve a model that better discriminates between patients who will or will not suffer from POD. Future work should investigate if less commonly used comorbidity indices are predictive of POD, or how either including different independent variables or using different models improves prediction of POD.

## Data Availability

All data used in the manuscript are freely available, upon completion of suitable training, from PhysioNet: https://physionet.org/content/mimiciv/3.1/.

